# Simplification of free-running cardiac magnetic resonance by respiratory phase using principal component analysis

**DOI:** 10.1101/2022.08.29.22279299

**Authors:** Ummul Afia Shammi, Zhijian Luan, Jia Xu, Aws Hamid, Lucia Flors, Joanne Cassani, Talissa A. Altes, Robert P. Thomen, Steven R. Van Doren

**Affiliations:** Department of Biomedical, Biological & Chemical Engineering, University of Missouri, Columbia, Missouri, United States of America; Institute for Data Science and Informatics, University of Missouri, Columbia, Missouri, United States of America; Department of Biochemistry, University of Missouri, Columbia, Missouri, United States of America; Department of Radiology, University of Missouri, Columbia, Missouri, United States of America

**Keywords:** real-time cardiac MRI, free-breathing MRI, motion correction, compressed sensing, post-processing, principal component analysis

## Abstract

Cardiac magnetic resonance imaging (CMR) provides many cardiac functional insights. The reliance of standard cine CMR upon breath holds is not feasible for some patients. Its process of combining multiple heartbeats is unsuited to arrhythmias. Real-time cine methods sidestep these problems but can introduce respiratory displacement of the heart. To aid CMR acquisitions during breathing, we developed post-processing software to diminish the effects of respiratory displacement of the heart. It uses principal component analysis to resolve respiratory motions from cardiac cycles in the dynamic image. The software groups heartbeats from expiration and inspiration to decrease the appearance of respiratory motion. The effects of respiratory motion and such motion correction were evaluated on short-axis views (acquired with compressed sensing) of 11 healthy subjects and 8 cardiac patients. The smallest correlation coefficients between end-systolic frames of the original dynamic scans averaged 0.79. After segregation of cardiac cycles by respiratory phase, the mean correlation coefficients between cardiac cycles were 0.94 ± 0.03 at end-expiration and 0.90 ± 0.08 at end-inspiration. The improvements in correlation coefficients were significant in paired t-tests, i.e., P ≤ 0.01 for healthy subjects and P ≤ 0.001 for heart patients at end-expiration. Two expert cardiothoracic radiologists, blinded to the processing, assessed the dynamic images in terms of blood-myocardial contrast, endocardial interface definition, and motion artifacts. Clinical assessment preferred cardiac cycles during end-expiration, which maintained or enhanced scores in 90% of healthy subjects and 83% of the heart patients. Performance remained high in a case of arrhythmia and irregular breathing. Heartbeats collected from end-expiration reliably mitigated respiratory motion when the new software was applied to DICOM files from real-time acquisitions.

## 1. Introduction

Cardiac magnetic resonance (CMR) is the gold standard for cardiac function evaluation [1,2]. It is a non-ionizing imaging modality that produces high spatial resolution images with excellent soft tissue contrast. Multi-slice 2D cine images can provide the anatomy of the entire heart and reveal the myocardial functionality of a patient in a single imaging session [3]. AHA/ACC guidelines strongly recommend CMR for patients with chest pain and either myopericarditis or aortic dissection [4]. In many settings calling for stress testing, the guidelines strongly recommended use of stress CMR [4].

Despite the appeal of cine CMR, it requires repeated breath holds and synchronization using electrocardiogram (ECG) to guide averaging and merging of frames into a composite cardiac cycle. These requirements can result in blurring, prolonged exams, and discomfort for patients with arrhythmias or who are unable to comply with the breath holds, e.g., due to chronic respiratory disease or unconsciousness in the ICU [5–7]. Heart failure, respiratory movement during breath holds, coughing, and patient movement due to discomfort inside the scanner commonly arise [8]. In addition, standard cine CMR does not capture the effects of the respiratory cycle upon cardiac cycles, i.e., heart rate increases with inspiration and decreases during expiration [9,10].

Various real-time acquisition methods can cross the barriers to application of cine MRI to patients who cannot comply with breath hold instructions or who are affected by an arrhythmia that interferes in cardiac gating [11–14]. For example, imaging of cardiac arrhythmias and abnormal wall motion was demonstrated with real-time CMR without breath holds [15–17]. It is also desirable to expedite imaging of children with congenital heart disease and decrease the need for sedating them [18,19]. The effects of percutaneous pulmonary valve implantation during exercise were studied with stress CMR in real-time [20]. However, the presence of breathing motion in real-time scans presents a challenge for the interpreting physician.

Several methods for fast acquisition of data have been developed. These fast image acquisition techniques can eliminate the need for hardware-dependent cardiac gating or respiratory navigation, allowing quick scanning of multiple tomographic views and instant image-based feedback [6,11]. Some of these acquisition techniques include parallel imaging [21–23] and sparse sampling by compressed sensing, radial, and spiral acquisitions [8,11,24–29]. Previously popularized pulse sequences include echo planer imaging (EPI) and inversion-recovery single-shot balanced steady-state free precession (BSSFP) [30–32]. These techniques improve spatial and temporal resolution. Real-time MRI acquisition of many frames throughout the range of respiratory motion introduces more variability among frames, posing challenges for motion correction. Image-based navigation and image registration and transformation schemes were introduced to correct respiratory motion [33,34]. However, image registration can distort images by forcing agreement with reference frames.

The work herein addresses, in part, the complexity added by real-time, free-breathing MRI acquired without cardiac gating. Our objectives were to accommodate the breathing of the subject and decrease the distraction of the ongoing breathing motion. We developed software for decreasing apparent respiratory motions in dynamic CMR, by grouping heartbeats according to respiratory phase. Fundamental to our strategy is to monitor the breathing motion retrospectively using post-processing, and then segregate the cardiac cycles collected during end-expiration and end-inspiration. We applied principal component analysis (PCA) to dissect the temporal fluctuations of dynamic series of CMR images (not in k-space). PCA is able to separate, both temporally and spatially, the large respiratory motions from cycles between systole and diastole, smaller cardiac motions, and turbulent blood flow [35,36]. Compared to segmentation, PCA and independent component analysis (ICA) hold inherent advantages in separation of concurrent respiratory and cardiac processes in the time dimension, globally throughout the dynamic 3D scan [35,37]. Compared to ICA, PCA is simpler to implement [35]. We exploited these advantages of PCA to characterize the phases of respiratory and cardiac cycles. We acquired real-time CMR scans during breathing without any gating, image-based navigators, or breath-holding maneuvers. From DICOM images with hundreds of frames, our new algorithm groups heartbeats by respiratory phase which quantifiably decreases apparent respiratory displacements of the heart in the set of beats. Not only does the appearance of respiratory motion decrease within the subsets of beats, but these subsets also retain diagnostic quality according to scores by two expert readers blinded to the post-processing. The algorithm made correct decisions when challenged by the arrhythmia and irregular breathing of a subject. The results suggest the potential utility of deriving cardiac cycles from end-expiration and perhaps end-inspiration from real-time CMR exams of patients unsuited for cine CMR.

## 2. Methods

### 2.1 Study subjects

Written informed consent was obtained from all subjects. Nineteen subjects (thirteen male, six female), aged from 24-73 years underwent free breathing cardiac MRI under an institutional review board protocol approved by the University of Missouri Institutional Review Board. The approved protocol is IRB #2007146 HS, named “Advancing Radiology Techniques” of Principal investigator Talissa Ann Altes, M.D. Eight of the subjects had previous cardiac history, while eleven were healthy controls. Demographic data of the subjects are given in Table 1.

**Table 1.** Demographic description of all subjects.

| Subject | Age range | Gender | Height [m] | Weight [kg] | Indication | Cardiac condition |
| --- | --- | --- | --- | --- | --- | --- |
| 1 | 18-49 | Female | 1.55 | 49.9 | Healthy | N/A |
| 2 | 18-49 | Male | 1.73 | 63.9 | Healthy | N/A |
| 3 | 18-49 | Male | 1.7 | 79.4 | Healthy | N/A |
| 4 | 18-49 | Female | 1.73 | 83.9 | Healthy | N/A |
| 5 | 50-80 | Male | 1.98 | 113.4 | Healthy | N/A |
| 6 | 50-80 | Female | 1.64 | 56.7 | Healthy | N/A |
| 7 | 50-80 | Male | 1.78 | 74.8 | Healthy | N/A |
| 8 | 50-80 | Female | 1.61 | 90.7 | Healthy | N/A |
| 9 | 50-80 | Female | 1.7 | 56.7 | Healthy | N/A |
| 10 | 18-49 | Male | 1.8 | 99.8 | Healthy | N/A |
| 11 | 50-80 | Male | 1.83 | 83.9 | Healthy | N/A |
| 12 | 50-80 | Female | 1.6 | 70.3 | Cardiac | Enlarged ventricle |
| 13 | 50-80 | Male | 1.88 | 95.2 | Cardiac | 1<sup>st</sup> degree AV block |
| 14 | 50-80 | Male | 1.8 | 97.1 | Cardiac | Blocked artery |
| 15 | 50-80 | Male | 1.75 | 95.2 | Cardiac | Diastolic heart failure; paroxysmal AFIB |
| 16 | 50-80 | Male | 1.8 | 79.4 | Cardiac | AFIB |
| 17 | 50-80 | Male | 1.83 | 108.8 | Cardiac | AFIB |
| 18 | 50-80 | Male | 1.83 | 106.6 | Cardiac | Paroxysmal AFIB; 1<sup>st</sup> degree AV block |
| 19 | 50-80 | Male | 1.79 | 125.6 | Cardiac | AFIB |

### 2.2 Imaging

The images were collected on a 3T Siemens Magnetom Vida (Siemens Healthcare, Erlangen, Germany) using an 18-channel chest radiofrequency coil and compressed sensing (CS, syngo MRXA20) to maximize both spatial and temporal resolution in short acquired during free breathing without gating or navigation. Real-time, free-breathing cardiac MR images were acquired with a conventional BSSFP sequence [31,32] and the Siemens implementation of compressed sensing. The rate of acquisition was between 12 and 22 frames/sec. The frame rate chosen by the Siemens software decreased with increased field of view if required for the subject. No averaging was used. Images were acquired with a 208 × 170 voxel matrix, in-plane isotropic voxel dimension of 1.4 - 1.73 mm, slice thickness of 6 mm, flip angle of 30°- 42°, TR of 43 - 96 ms and a total acquisition time of 22-35 sec.

### 2.3 Algorithm for simplification of real-time MRI

Our algorithm simplifies real-time free-breathing MRI scan by grouping heartbeats according to respiratory phase. The framework for grouping heartbeat according to respiratory phase is shown as a block diagram in Fig 1 and described below. The algorithm employs and automates three major steps:

**Fig. 1.**
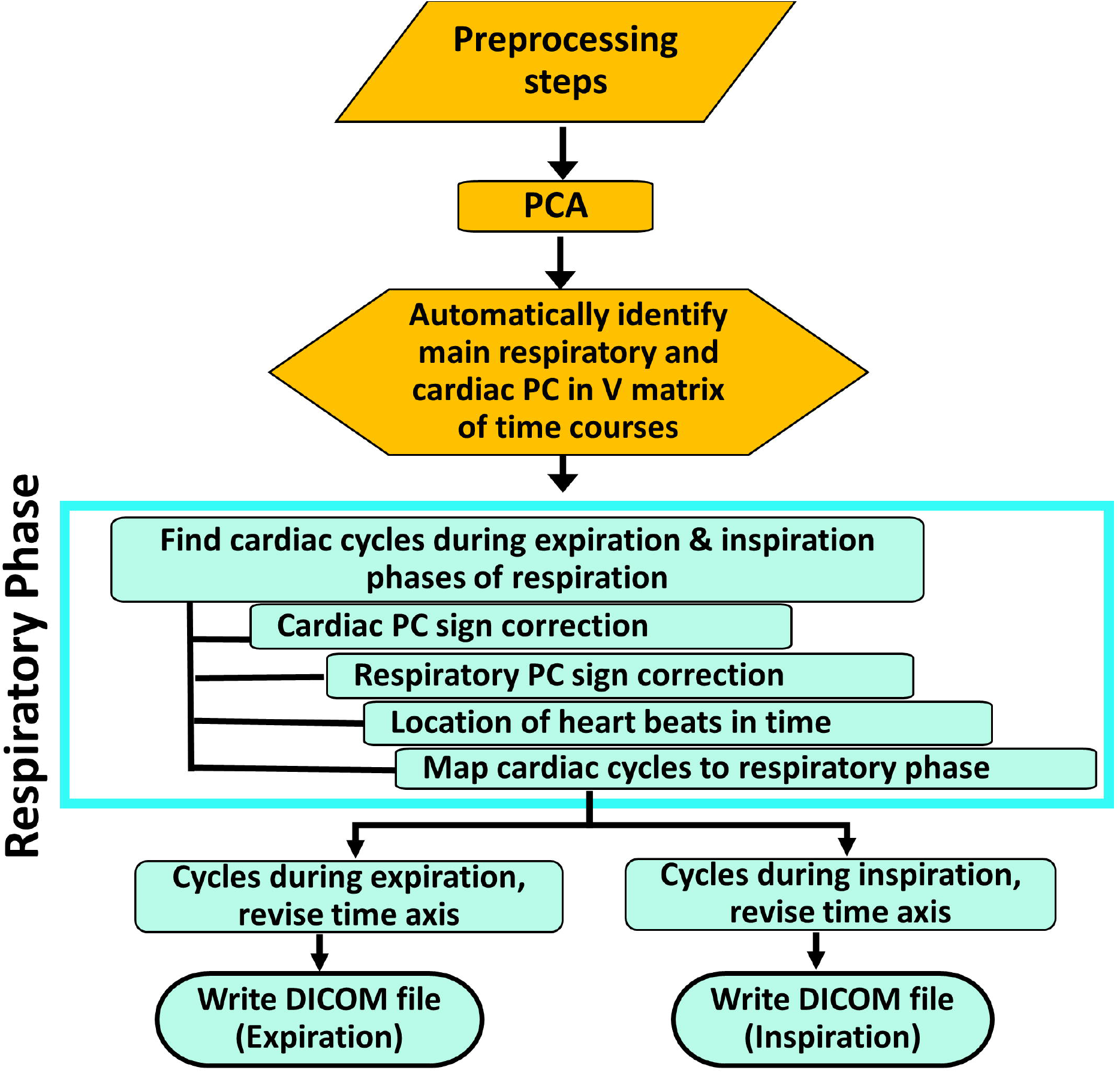
Algorithm developed to group heartbeats by respiratory phase.

1. Preprocess to resolve principal components (PCs) and their time courses with potential to report on respiratory or cardiac contractions.
2. Identify cardiac cycles during breaths out and breaths in.
3. Create DICOM files that collect and segregate cardiac cycles at end-expiration and end-inspiration.

The preprocessing strategy (S1 Fig) was detailed previously [35]. Identification, selection, and isolation of cardiac cycles at end-expiration and end-inspiration of from dynamic CMR images (Fig 1) have been developed herein.

#### 2.3.1 Preprocessing: Implementation of PCA using TREND software

The approach taken requires resolution of the largest of the time-dependent signals present in CMR scans acquired during breathing in real-time. Preprocessing supplied these candidate signals. Principal component analysis (PCA) reduces the dimensionality of data with many experimental variables [38]. A new software platform called TRENDimage (version 1.9.7.1), developed from TREND [35], was used in order to perform unfold PCA upon stacks of DICOM image frames, captured either in a single file in enhanced DICOM format or in a directory of hundreds of individual frames (Fig 1). The first preprocessing step of TREND “unfolds” the 3D scan into a 2D data matrix wherein each 2D image frame is cast into a 1D column (Fig S1 in the Supplemental Material). The time dimension remains unaltered by this process. The resulting 2D data matrix is then mean-centered (i.e., the mean is subtracted from each element of the matrix) prior to the singular value decomposition step of PCA that decomposes the preprocessed matrix X’ into three matrices [35]:

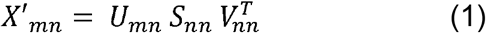

Here, *m* refers to the number of rows and n to the points per row. *U*_*mn*_ and 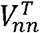 matrices are orthogonal matrices and *S*_*mn*_ is a diagonal matrix. *V*_*nn*_ contains the trends of change in motion over time, representing the principal components (PC) of that image series. The workflow of the original TREND software is summarized in S1 Fig.

#### 2.3.2 Identification of cardiac cycles and their locations during respiration

##### 2.3.2.1 Automatic identification of main respiratory and cardiac time courses

Since ECG and respiratory navigation were avoided during image acquisition, we obtained the time courses of the cardiac cycles and respiratory cycles retrospectively from PCA instead (Fig 2). After resolving dozens of motional processes into principal components (PCs) using PCA, the next step was to identify principal respiratory and cardiac PCs. Fig 2A shows time courses of the four largest PCs obtained from a real-time CMR scan. We distinguished respiratory rates from heart rates in the power spectra (Fourier transforms) of the oscillations of the PCs in real time (Fig 2B).

**Fig. 2.**
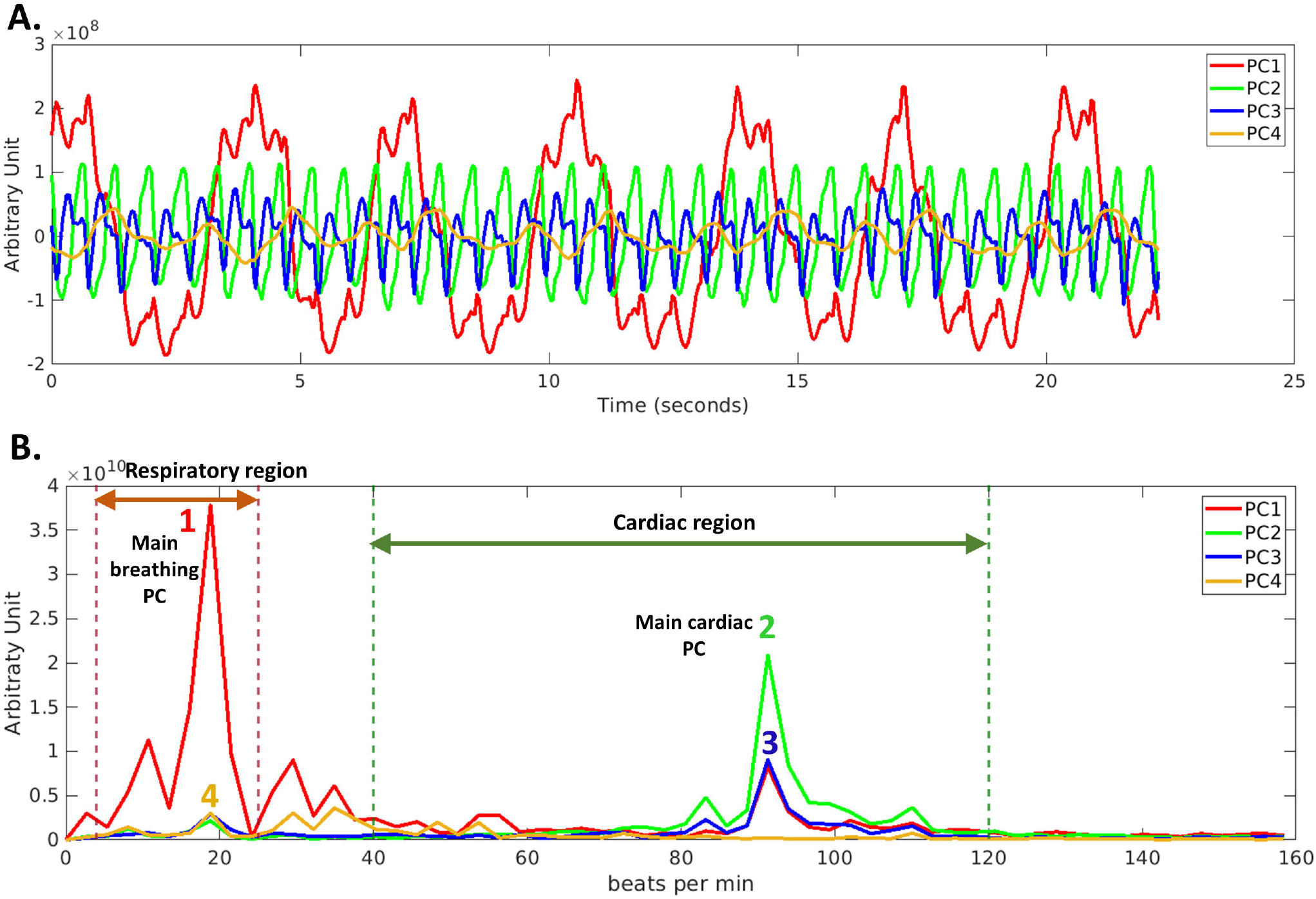
Evaluation of time courses of major principal components for oscillations at respiratory and heart rates. (A) Plots of individual PC values vs. time of a three-chamber slice scan during free breathing. (B) Magnitude Fourier transforms of the time courses from panel A clarify the frequencies of the fluctuations.

##### 2.3.2.2 Automatic recognition of cardiac cycles

Since an ECG signal was not acquired, we obtained the time courses of the cardiac cycles retrospectively from PCA instead (Fig 2). PCs are arbitrary in sign [35]. Consequently, we defined systole as positive in sign, based on the lesser brightness of the smaller volume of blood than during diastole (S2 Fig). The most distinct and consistently identifiable starting point of each cycle in such time courses is at end-systole because it is narrower and more distinct than end-diastole (Fig 2A). A complete cardiac cycle was defined from end-systole to end-systole. To define complete cardiac cycles, the algorithm looked for the cardiac time course passing through an amplitude of zero (S2 Fig). For better estimates of the point in time of each peak at end-systole, cubic interpolation boosted the number of time points by 100-fold for peak picking. If two or three peaks were candidate peaks at end-systole, the tallest peak was chosen to mark end-systole. False, smaller peaks were filtered out when (i) shorter than a smoothed trend line through the cardiac time course generated by low-pass filtering or (ii) when within 0.4 sec of a taller peak.

Potentially overlooked cardiac cycles and missing peaks at end-systole were sought in a second search. In cases of cardiac cycles that appeared longer than the mean length of a heartbeat by > 1.5 σ, the follow-up search lowered the threshold for peak picking to detect weaker nearby peaks at end-systole. The lower threshold was set at the height of the weakest end-systole peak found above a trendline through the cardiac time course, smoothed by low-pass-filtering.

#### 2.3.3 DICOM files of cardiac cycles from expiration and inspiration automatically

Finally, the algorithm accumulated all the heartbeats found during expiration and inspiration (S3-S4 Figs) and creates two separate DICOM files. The resultant files were used for statistical analysis and assessment by cardiothoracic radiologists.

#### 2.3.4 Coding environments

Algorithms to analyze PCs and their time courses were initially developed in MatLab version 2018b (MathWorks, Natick, MA). The algorithms were then ported to Python 3.7. A standalone version (not requiring a Python compiler) has been prepared for simple “two-click” operation under Windows.

#### 2.3.5 Operation of the new software for motion correction

The simple step-by-step operation of the software is described at <u>dx.doi.org/10.17504/protocols.io.36wgq72j5vk5/v1</u> under DOI: dx.doi.org/10.17504/protocols.io.36wgq72j5vk5/v1. Please contact Steven Van Doren and Brett Maland regarding licensing and pricing of the software.

### 2.4 Evaluations

#### 2.4.5 Statistical analysis

2D correlation coefficients were calculated between corresponding voxel signal intensities of each frame at end-systole with a single reference frame at end-systole from the beginning of the scan. For plots and paired-sample t-tests, the lowest correlation coefficient was selected from the set of correlation coefficients between frames. This was repeated before and after motion correction by respiratory phase for assessment of the reproducibility of frames at end-systole. The mean as well as 25^th^ and 75^th^ percentiles of the lowest correlation coefficient for sets of scans were visualized using a box plot. We also calculated the root-mean-squared deviation of the centers-of-brightness or centroids [39] within the heart. We displayed distributions of centroid values with box plots. A paired samples t-test was performed for each evaluation metric before and after motion correction. P-values less than 0.05 were considered significant. Displacement field vectors were used to visualize local motions before and after motion correction.

#### 2.4.2 Expert scoring of cardiac imaging

We presented images with and without retrospective motion correction to two expert cardiothoracic radiologists blinded to the post-processing. They evaluated the CMR quality in terms of blood-myocardial contrast, endocardial interface definition, and motion artifact. Each criterion was graded on a scale of 1 (insufficient) to 5 (excellent) (Table 2) on review of short axis slices.

**Table 2.**
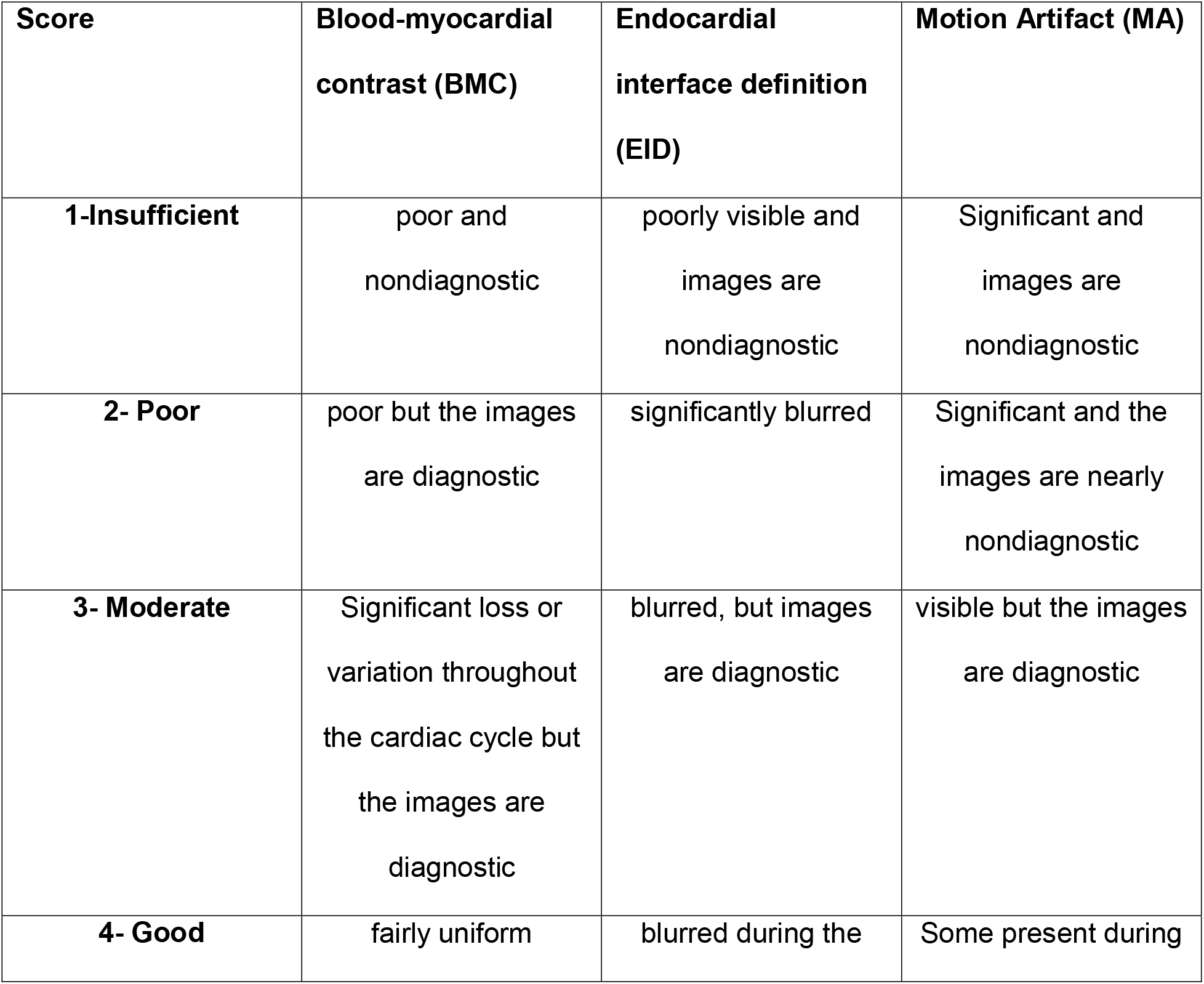
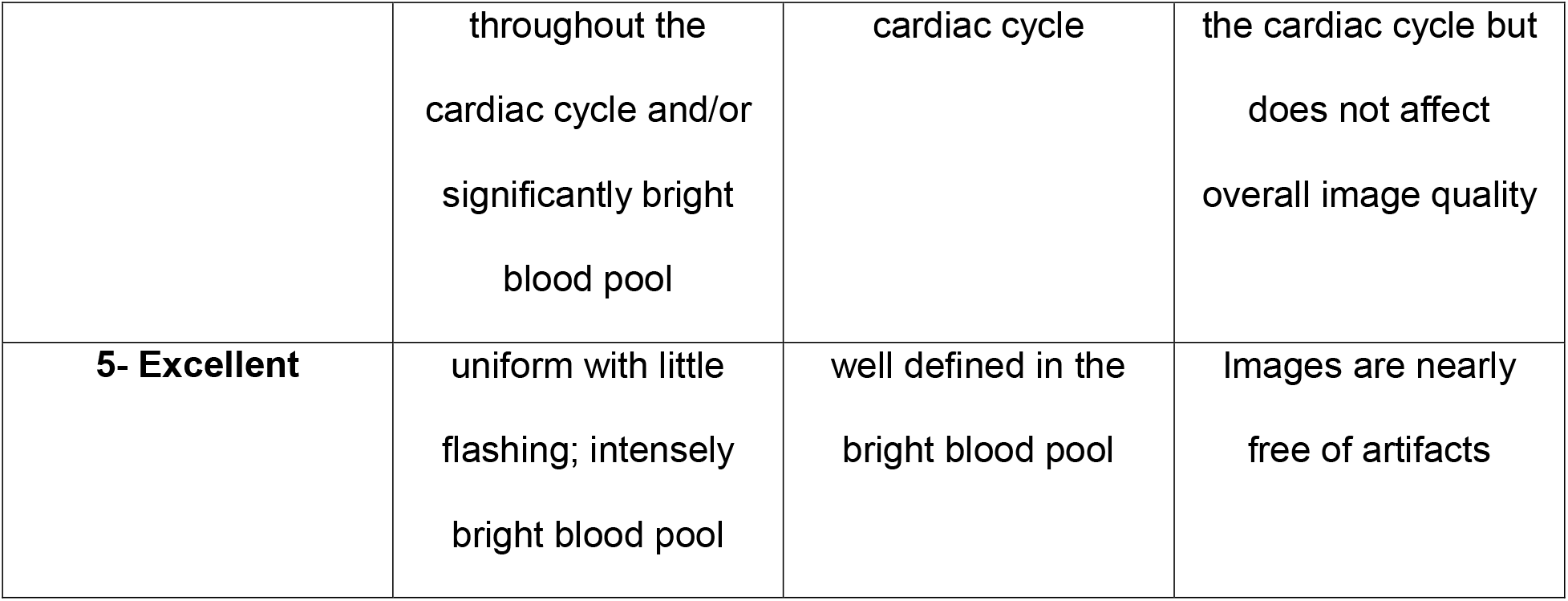
Criteria used by cardiothoracic radiologists in scoring the images.

## 3. Results

### 3.1 Resolution of cardiac cycles and phases

The new algorithm identified the main principal components (PCs) at respiratory and cardiac contractile rates from power spectra of the PCs (Fig 2). This achievement required optimization of the threshold by which the peak height at the respiratory rate was required to exceed the peak height at the heart rate, and vice versa (Fig 2B). A ratio of 2.2 emerged from the optimization. The algorithm accurately corrected the sign of the respiratory PC in 93% of transverse and long-axis scans. An error interchanged the labels on the expiratory and inspiratory subsets of cardiac cycles derived from the original scan. The algorithm corrected the sign of the cardiac PC accurately in 90% of transverse and long-axis scans. An error resulted in cardiac cycles being marked from end-diastole to end-diastole rather than end-systole to end-systole.

### 3.2 Suppression of apparent breathing motion

Suppression of respiratory motion in the background was validated by evaluating impact on movement of the heart (Figs. 3, 4), reproducibility of frames from cardiac cycles (Fig 5), and clinical assessments of the dynamic images (Fig 6).

**Fig. 3.**
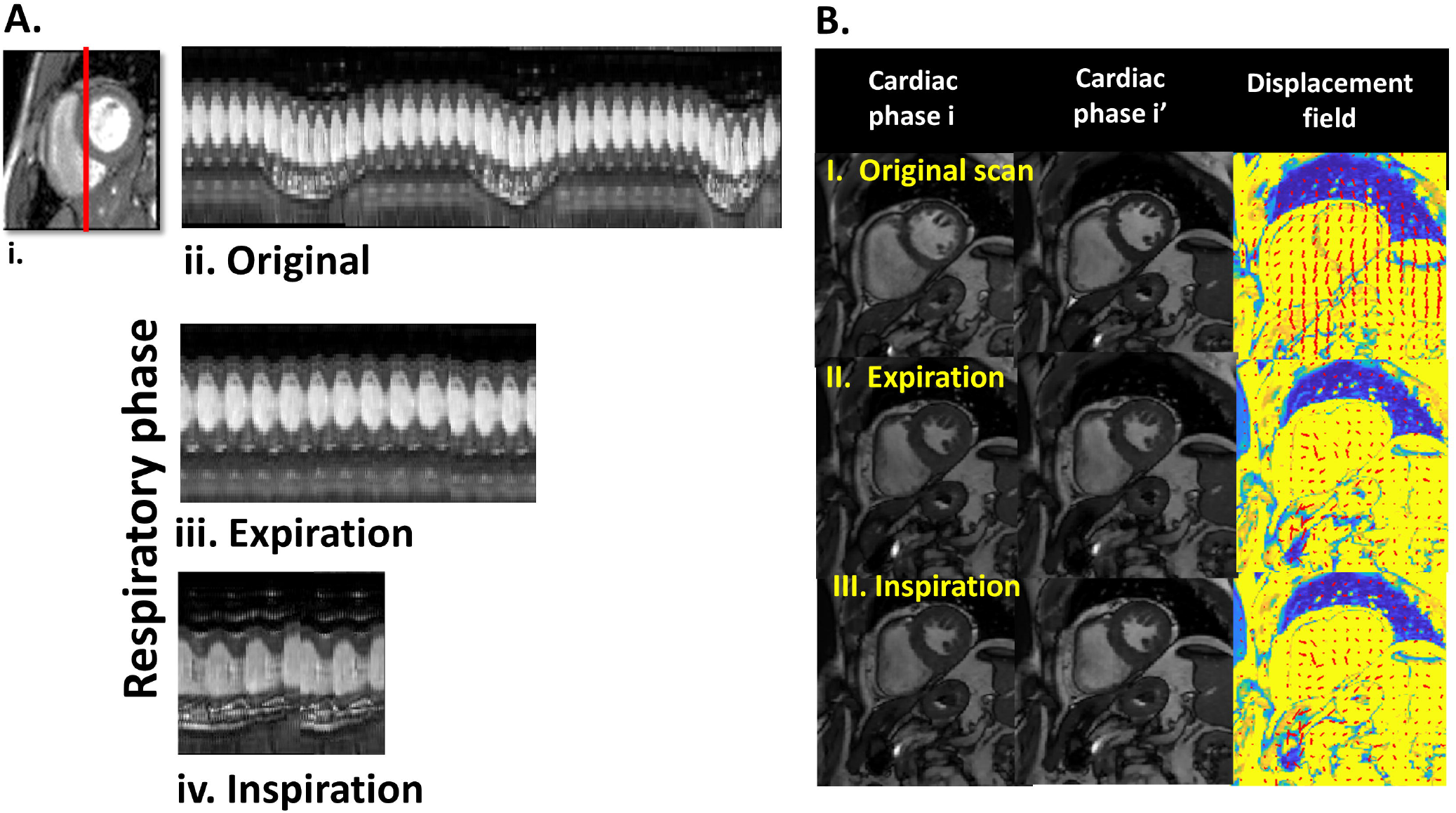
Respiratory displacement of heart in transverse slice mitigated by respiratory phase. (A) A column of pixels (shown in red vertical line) is plotted from frames before (ii) and after grouping by respiratory phase (iii, iv) in a SAX view (B) Displacement field vectors of the SAX scan before and after this motion correction are less affected by respiratory displacement of the heart. The displacement fields were derived from the pairs of frames (i and i’) at matched cardiac phases but at distinct respiratory phases in the top row.

**Fig. 4.**
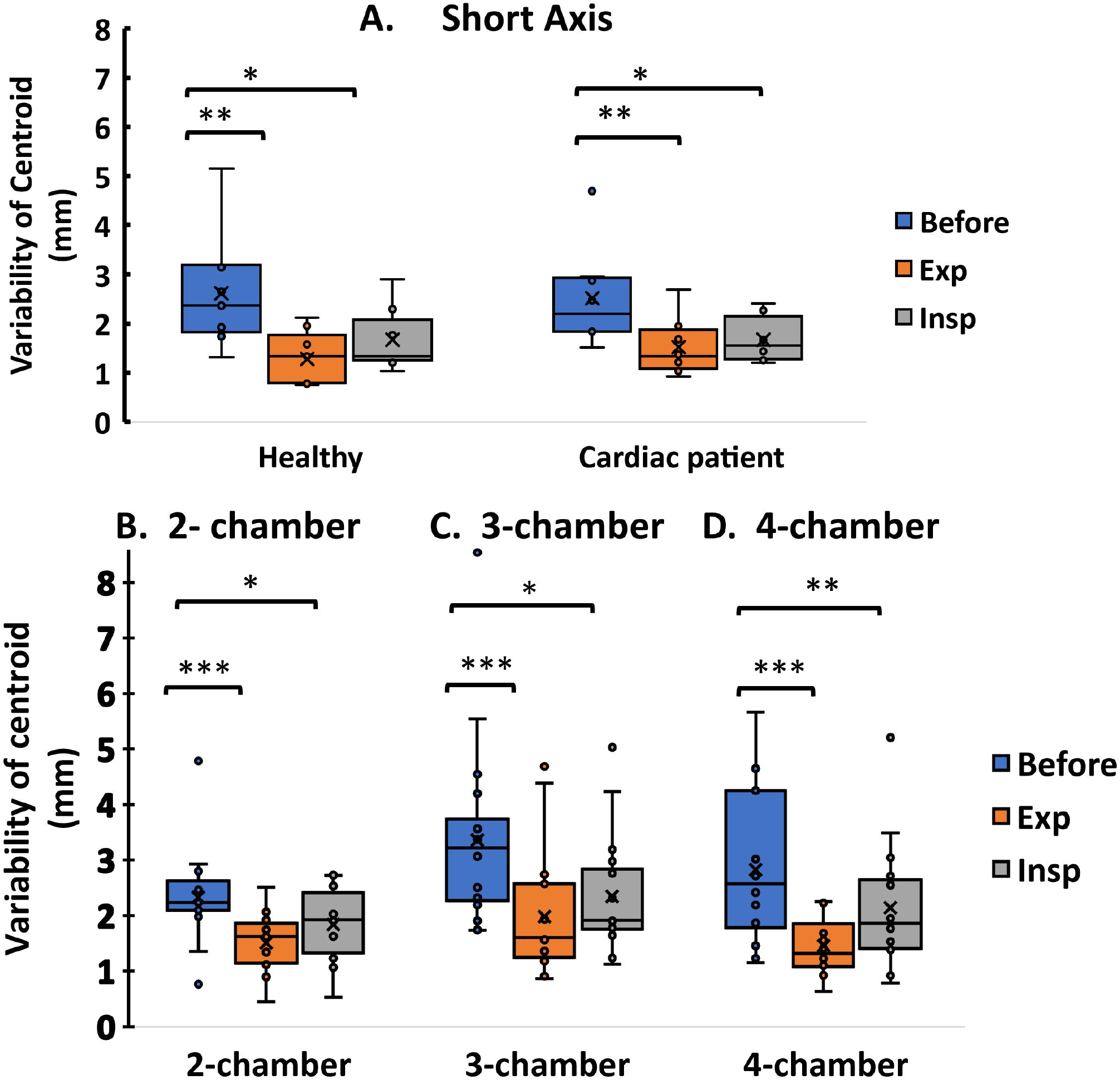
Variability of the centroid of real-time, dynamic CMRI scans, before and after grouping by respiratory phase. Results for short-axis views are shown in (A). Results for long-axis views are shown in the lower row for 2-chamber views (B), 3-chamber views (C), and 4-chamber views (D). The centroid is centered on the bright blood in the heart. Paired t-test results with P ≤ 0.05, P ≤ 0.01, or P ≤ 0.001 are represented by *, **, or ***, respectively.

**Fig. 5.**
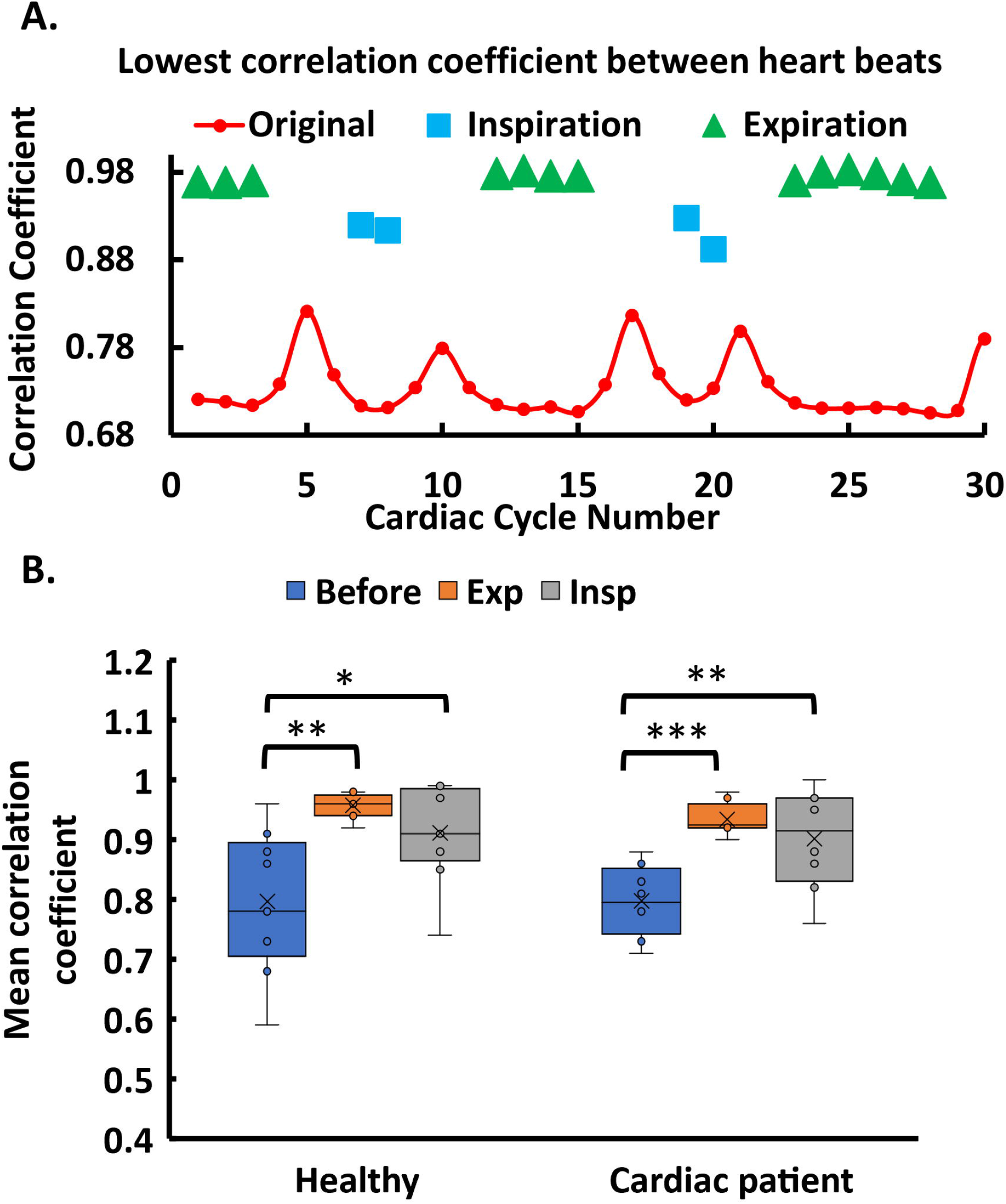
Image consistency between heartbeats is improved by grouping the beats from end-expiration or end-inspiration. Correlation coefficients were calculated among the end-systolic frames of every heartbeat. Panel (A) shows one example of a short axis view. Panel (B) summarize results for all SAX scans (for healthy controls and cardiac patients). Each point represents the average lowest correlation coefficient for a scan before and after grouping by respiratory phase. Each box plot shows the median as a horizontal line and the 25th and 75th percentiles as the edges of the box. Statistical significance with P ≤ 0.05, P ≤ 0.01, or P ≤ 0.001 is indicated by *, **, or ***, respectively.

**Fig. 6.**
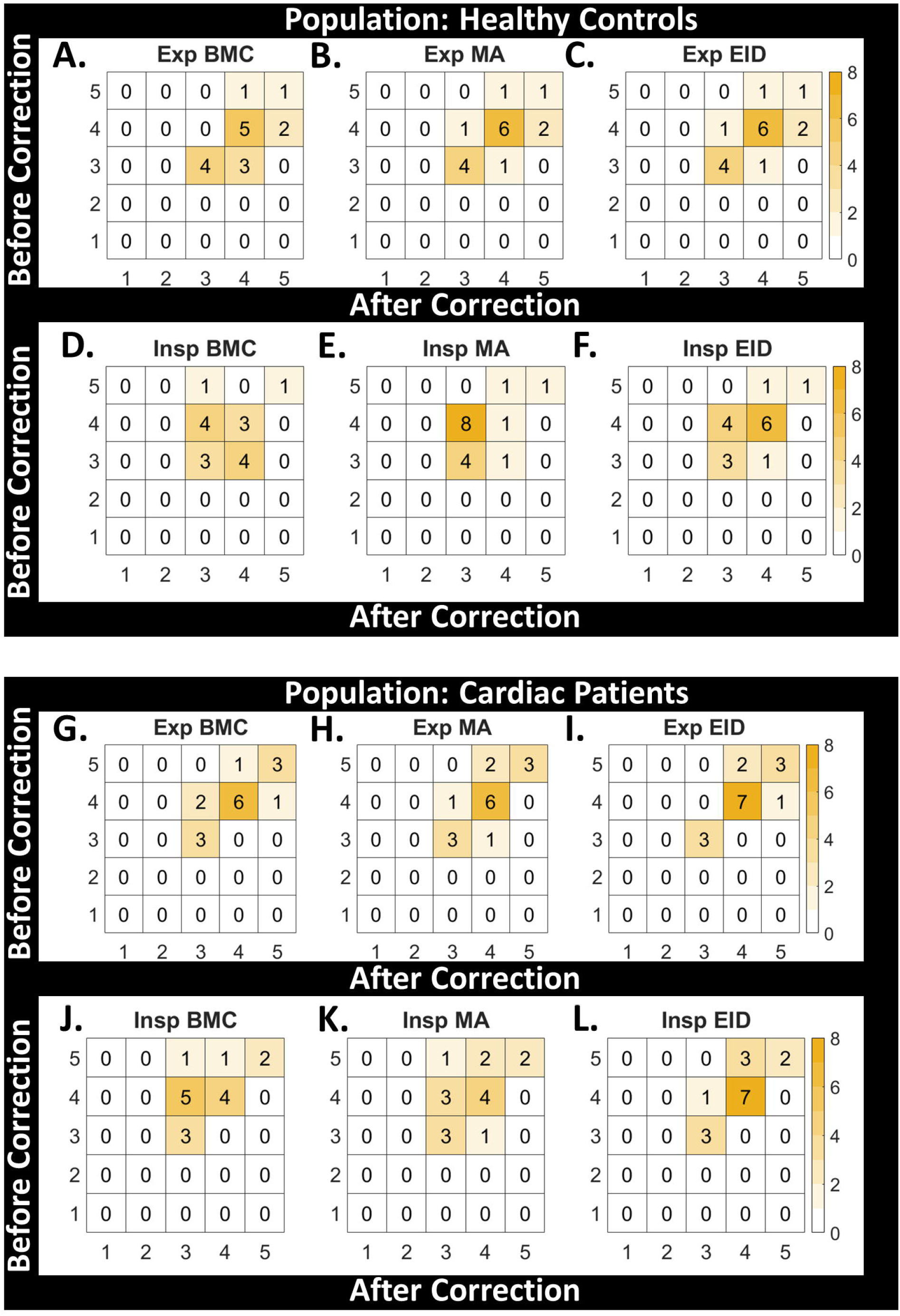
Scores of image quality of short axis views of volunteers, before and after grouping by respiratory phase, by two expert radiologists blinded to the post-processing used. (A-F) Scores for the healthy volunteers are tabulated. (G-L) Scores for volunteers with cardiac symptoms. (A-C, G-I) These comparisons tally the scores before and after collecting beats from expiration. (D-F, J-L) These comparisons tally the scores before and after collecting beats from inspiration. (A,D,G,J) Blood-myocardial contrast. (B,E,H,K) Motion artifact. (C,F,I,L) Endocardial interface definition.

#### 3.2.1 Evidence of less respiratory motion

The management of breathing motion by respiratory phase can be visually evaluated by comparison of a column of pixels from each frame before and after segregation into cardiac cycles at end-expiration or end-inspiration (Fig 3A). Among the columns from cardiac cycles segregated by respiratory phase, there is less respiratory displacement of the heart (Fig 3A). The suppression of large breathing motion was substantial during end-inspiration and excellent during end-expiration. Displacement of the heart between frames at differing respiratory phase, but similar cardiac phase, is shown in Fig 3B. The displacement fields of the short-axis scans acquired during free breathing are dominated by respiratory movement (right panel of Fig 3B I). After automatic grouping by respiratory phase, the displacement fields are dominated instead by cardiac motions (right panels of Fig 3B II, III).

Respiratory movement of the heart was monitored in all acquired scans using the center-of-brightness, centered upon the blood in the heart (Fig 4). Inspiratory groups of heartbeats for both transverse and long-axis views had less respiratory motion than the original scans (P ≤ 0.05; Fig 4). Expiratory groups of heart beats enjoyed more significant decreases of respiratory motions, both for short-axis views (P ≤ 0.01) and long-axis views (P ≤ 0.001; Fig 4). Thus, cardiac cycles from expiration were improved even more than those from inspiration.

#### 3.2.2 Reproducibility of heartbeats

The uniformity and reproducibility of the heartbeats was assessed using correlation coefficients between voxel signal intensities of each frame at end-systole with a reference frame at end-systole, both before and after motion correction. The smallest of those correlation coefficients are plotted in the example in Fig 5A. This measure of the consistency of the heartbeats was compared by t-tests before and after correction of short axis view of the 19 subjects.

Motion-corrected images rendered the heartbeats clearly more consistent with one another (Fig 5B). The improvements by respiratory grouping of heartbeats are statistically significant. The inspiratory groups of heartbeats improved the correlation coefficients with *P* ≤ 0.05 for the healthy subjects and *P* ≤ 0.01 for the subjects with histories of cardiac symptoms (Fig 5B). The expiratory groups improved the correlation coefficients with *P* ≤ 0.01 for the healthy subjects and *P* ≤ 0.001 for the subjects with a history of cardiac symptoms. See Supplementary videos S1, S2 (short-axis view) and S3 (a long-axis view) for examples of scans of subjects without and with segregation of cardiac cycles. The hypothesis that grouping by respiratory phase not only improved the correlation coefficients but also preserved the clarity of cardiac structure was tested by clinical scoring.

#### 3.2.3 Image evaluations by expert readers

The qualities of the motion-corrected short-axis images from the cohort of volunteers were compared alongside the original images in blinded fashion by two expert cardiothoracic radiologists. The expert readers scored the image quality by criteria of blood-myocardial contrast, endocardial interface definition, and motion artifact (Table 2). The scores of image qualities of healthy subjects and heart patients before and after motion correction are shown in Fig 6 by heatmaps representing the combined opinions of the two radiologists. Among the healthy subjects, the blinded scores across the three criteria were judged the same or better in 90% of the evaluations of images of cardiac cycles from expiration and in 60% of the evaluations of those from inspiration (Figs 6A-F). Among the heart patients, the blinded scores were judged the same or better in 83% of images of cardiac cycles from expiration and 65% of those from inspiration (Fig 6G-L). However, the residual vertical displacement of the heart by inspiration increased the perceived motion artifact of cardiac cycles collected from inspiration in 56% of the healthy subjects and 36% of the heart patients (Fig 6E, K). This perception could result from the large and varied vertical heart displacement in the inspiratory movies (Supplementary videos S1, S2). Nonetheless, the image quality was preserved or even enhanced in 86% of all images of cardiac cycles from expiration (Fig 6).

### 3.3 A case of non-uniform respiratory and cardiac cycles

The performance of the grouping algorithm was scrutinized carefully on the challenging case of a subject with an arrythmia and non-uniform breathing. The algorithm worked fine in all of the essential steps: The sign corrections of the time courses of the cardiac and respiratory PCs were accurate. Consequently, the software correctly assigned the inspiratory and expiratory phases in the respiratory time course and the systolic and diastolic phases in the cardiac time courses (Fig 7). The software accurately divided the heartbeats from end-systole to end-systole despite the varying lengths of the cardiac cycles (Fig 7B). Notably, the PCA-based division of the cardiac cycles identified heartbeats of unusual length, i.e., pairs in which a short beat was followed by a long beat (see yellow highlights in Fig 7 and Supplementary video S4).

**Fig. 7.**
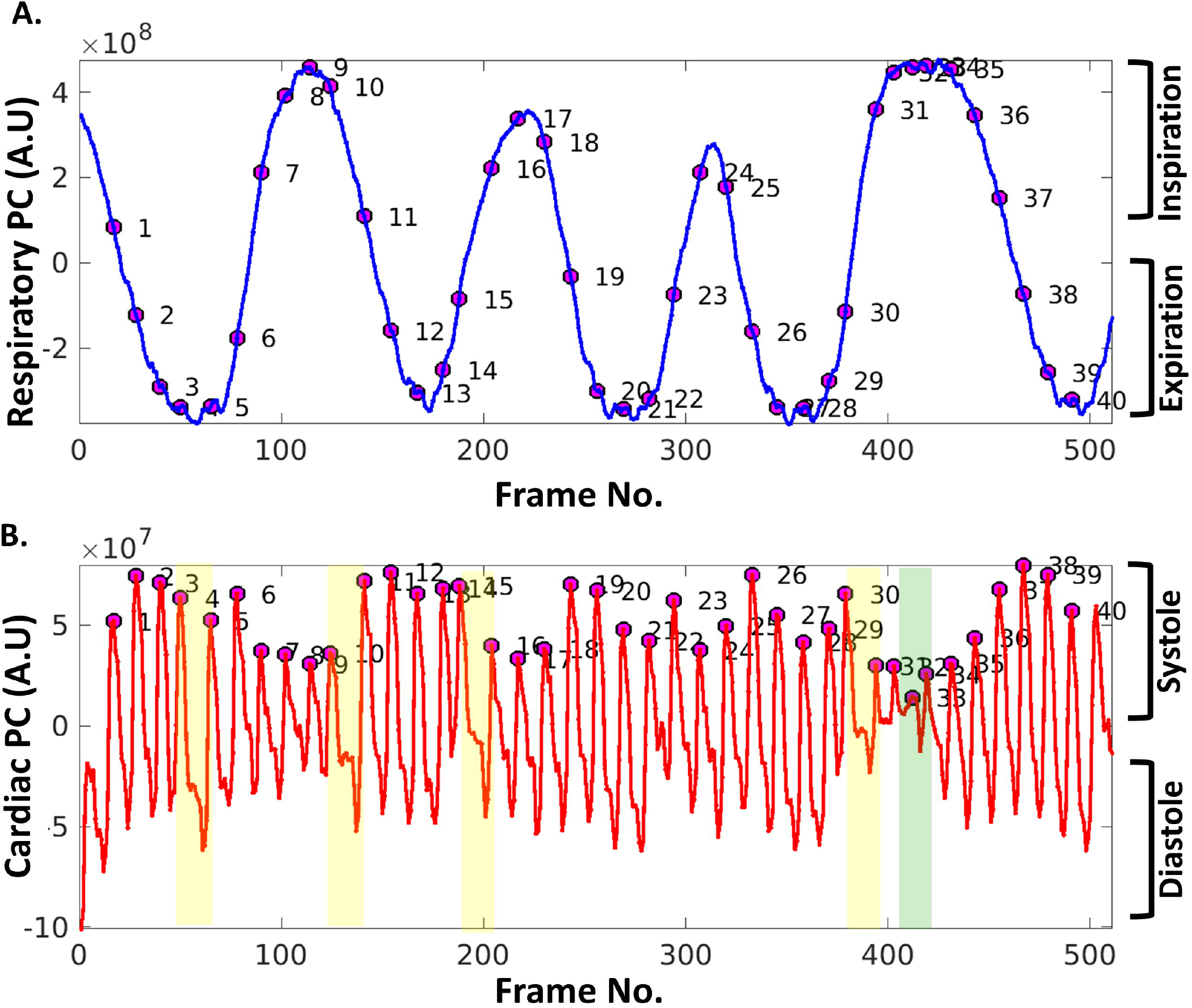
Time courses of respiratory and cardiac PCs of a subject with an arrythmia. (A) Correct identification of the respiratory phases was confirmed by viewing the DICOM file of this male subject (between 50 and 80 years old). (B) The cardiac cycles are divided end-systole to end-systole in the time course of the cardiac PC. Beats 4, 10, 15, and 30 were longer than usual (highlighted by yellow), and each followed a shorter-than-average beat. Beats 32 and 33 (highlighted by green) were shorter than usual.

## 4 Discussion

### 4.1 Technical advances

We sought to simplify CMR scans collected during breathing in real-time without cardiac gating. Our objectives were to accommodate the breathing of the subject and decrease the distracting appearance of the breathing motion. Our new software decreased the apparent heart displacement by breathing, relative to other cardiac cycles of similar respiratory phase. The algorithm exploits the advantages of PCA in both *temporal* and spatial resolution throughout the data matrix, without any training involved in this resolution [35,36]. The resulting collections of heartbeats from expiration are mostly free of distracting respiratory motion. The heartbeats segregated by respiratory phase may enhance the apparent blood-myocardial contrast and endocardial edge delineation of some short-axis scans acquired during breathing. When faced with irregular heart contractions and irregular breathing, this software that utilizes PCA worked well in creating the expiratory and inspiratory portions of the original scan.

The ICA alternative to PCA offers the advantages of accommodating non-Gaussian data distributions [40], superior separation of signal components by the Combi ICA implementation, and a history of successful application to fMRI in resolving blood flow signals in the brain [37,41]. However, ICA was inconvenient for the automation needed for this study because (i) the independent components (ICs) generated must be fewer or equal to the number of true components to be correct [42], (ii) the ICs are unordered and require sorting, and (iii) the best ICA run needs to be selected from multiple ICA calculations [43].

### 4.2 Anticipated relevance to patients

Real-time cardiac MRI, acquired rapidly by a variety of innovative methods, provides reliably high image quality [11]. By foregoing averaging, patients with an irregular heartbeat can be imaged, in principle. Other patients who are less compliant to averaged CMR might benefit by imaging during breathing. Such patients may include those suffering dyspnea from COPD or asthma or those in the ICU with a traumatic brain injury. Mitigation of apparent respiratory motion in scans acquired during breathing, by this post-processing software or other methods, offer potential to relax expectations of regular heart rhythm, regular respiration, and breath holds. To the extent that real-time acquisitions might supplant breath-held parts of CMR exams, exams could become shorter and more tolerable for non-compliant patients, while enhancing cost efficiency for clinics.

### 4.3 Limitations

An earlier version of the Siemens compressed sensing package acquired the real-time CMR scans of young adult volunteers at 22 frames/sec. However, software upgrades and subjects requiring larger field of view resulted in slower frame rates. Images from the slower acquisitions were less crisp and had lower temporal resolution. While our algorithms manage normal free breathing, large inspirations cause large vertical displacements of the heart. This affects motion-corrected inspiratory heartbeats and introduces large through-plane motion to long-axis views. Inspirations of differing amplitude result in different positions of the diaphragm. This results in different respiratory shifts of the heart when comparing heartbeats from different end-inspiratory periods of the scan. To compensate for this limitation, subjects could be asked to breathe more shallowly during imaging. Lastly, our study was limited to small numbers of volunteers who were healthy or with a cardiac condition. Effects on assessment of cardiac function remain to be investigated.

## 5. Conclusions

We simplified the free-breathing, dynamic cardiac MRI images of small numbers of healthy and symptomatic subjects. This was achieved by segregating cardiac cycles from end-expiration and end-inspiration phases of respiration by post-processing. The technical strategy is novel and distinct from methods reliant upon image registration. The post-processing strategy is vendor-neutral and neutral to the type of real-time image acquisition. The software could merit trial with a broader range of real-time image acquisitions. The combination of real-time acquisitions with this post-processing software offers the potential to facilitate CMR scanning of patients non-compliant to cine protocols. This includes patients too frail for multiple breath holds and patients with arrhythmias unsuitable for averaging. The simplification of free-running CMR by respiratory phase fosters possibilities of visualizing the variability of cardiac cycles with respiratory phase and arrhythmias. Despite residual motion artifacts during deep breaths in, inspiratory groups of cardiac cycles might reveal effects of inspiration upon cardiac behavior. The image quality of the cardiac cycles from expiration is superior, and attractive for wider evaluation.

## Data Availability

The results generated for each human subject (anonymized) are available at the OSF
repository at https://osf.io/4unbe/ under the DOI 10.17605/OSF.IO/4UNBE. The data available include real-time cardiac MRI scans before and after motion correction by respiratory phase (.avi movie format). Plots of the time courses of respiration, heart contraction, and improvement in correlation coefficients are also available for each
subject. A protocol describing step-by-step operation of the new motion correction software is
available at dx.doi.org/10.17504/protocols.io.36wgq72j5vk5/v1 under DOI: dx.doi.org/10.17504/protocols.io.36wgq72j5vk5/v
Academic licensing of the software developed can be requested by contacting the corresponding author and the Office of Technology Advancement at the University of Missouri (contact Brett Maland, email:
Tel: 1-573-882-1046, W1030 Lafferre Hall, Univ. of Missouri, Columbia MO 65211 USA). Commercial licenses are also available. To request a quote, contact Steven Van Doren or Brett Maland.

## Abbreviations

BMC: Blood-myocardial contrast
CC: Pearson correlation coefficient
CMR: Cardiovascular magnetic resonance
COPD: Chronic obstructive pulmonary disease
ECG: Electrocardiography
EID: Endocardial interface definition
EPI: Echo planner imaging
BSSFP: Balanced steady-state free precession
IC: independent component
ICA: Independent component analysis
ICU: Intensive care unit
IRB: Institutional review board
MA: Motion artifact
MoCo: Motion correction
PC: Principal component
PCA: Principal component analysis
RF: Radiofrequency
RMSD: Root mean square deviation
SAX: short axis view
SD: standard deviation
TREND: Tracking Equilibrium and Nonequilibrium Shifts in Data
VD: Vertical distance

## Acknowledgment

We thank Mark Burton for regular assistance in setting up image acquisition.

## Supporting Information

**S1 Fig. Workflow using the original TREND software for user-chosen motion correction**. The core matrix processing of the TREND software available at beginning of this study is highlighted in the gray box. Xu and Van Doren (2017, DOI: <u>10.1016/j.bpj.2016.12.018</u>) illustrated the ability of the *user* to decide and suppress breathing motion manually (cyan box) in images in .avi and .mp4 movie formats.

**S2 Fig. Cardiac PC sign correction**. Main cardiac time courses from (a) a 2-chamber view and (b) a 3-chamber view from PCA are plotted. At diastole, blood flows into the heart causing the image to be brighter. At systole, the ejection of blood from the heart causes the image to be darker. The brightness provided the basis to correct the arbitrary sign from PCA.

**S3 Fig. Respiratory PC sign correction associates end-expiration with more heart beats than end-inspiration**. More local peaks on the negative side of the curve lie in the end-expiration region. End-inspiration is set to be positive.

**S4 Fig. Mapping of heart beats to end-inspiration and end-expiration phases of the respiratory cycle represented by the main breathing PC**. Gold highlights heart beats confined to end-inspiration while blue marks beat in end-expiration.

**S5 Fig. Scores of image quality of the volunteers by reader**. Two expert radiologists scored the images in the short-axis view, while they were blinded to the post-processing used.

**S1 Video Sax Healthy Control.mp4**

**S2 Video Sax Cardiac Patient.mp4**

**S3 Video 2ch Healthy Control.mp4**

**S4 Video PC Analysis of Cardiac Patient.mp4**

**Supporting Protocol** CMR_MoCo_software_operation_protocol.pdf

## References

1. Constantine G, Shan K, Flamm SD, Sivananthan MU. Role of MRI in clinical cardiology. Lancet. 2004;363: 2162–2171. doi:https://doi.org/10.1016/S0140-6736(04)16509-4

2. Kramer CM, Barkhausen J, Bucciarelli-Ducci C, Flamm SD, Kim RJ, Nagel E. Standardized cardiovascular magnetic resonance imaging (CMR) protocols: 2020 update. J Cardiovasc Magn Reson. 2020;22: 17. doi:10.1186/s12968-020-00607-1

3. Barkhausen J, Goyen M, Rühm SG, Eggebrecht H, Debatin JF, Ladd ME. Assessment of Ventricular Function with Single Breath-Hold Real-Time Steady-State Free Precession Cine MR Imaging. Am J Roentgenol. 2002;178: 731–735. doi:10.2214/ajr.178.3.1780731

4. Gulati M, Levy PD, Mukherjee D, Amsterdam E, Bhatt DL, Birtcher KK, et al. 2021 AHA/ACC/ASE/CHEST/SAEM/SCCT/SCMR Guideline for the Evaluation and Diagnosis of Chest Pain. J Am Coll Cardiol. 2021;78: e187–e285. doi:10.1016/j.jacc.2021.07.053

5. Zhang S, Joseph AA, Voit D, Schaetz S, Merboldt K-D, Unterberg-Buchwald C, et al. Real-time magnetic resonance imaging of cardiac function and flow—Recent progress. Quant Imaging Med Surg. 2014;4: 313–329. Available: http://qims.amegroups.com/article/view/4050

6. C YP, B KA, C LA, H LD, Chris H H MC, et al. New real-time interactive cardiac magnetic resonance imaging system complements echocardiography. J Am Coll Cardiol. 1998;32: 2049–2056. doi:10.1016/S0735-1097(98)00462-8

7. Barnett LA, Prior JA, Kadam UT, Jordan KP. Chest pain and shortness of breath in cardiovascular disease: a prospective cohort study in UK primary care. BMJ Open. 2017;7. doi:10.1136/bmjopen-2017-015857

8. Usman M, Atkinson D, Odille F, Kolbitsch C, Vaillant G, Schaeffter T, et al. Motion corrected compressed sensing for free-breathing dynamic cardiac MRI. Magn Reson Med. 2013;70: 504–516. doi:10.1002/mrm.24463

9. Strauss-Blasche G, Moser M, Voica M, McLeod D, Klammer N, Marktl W. Relative Timing Of Inspiration And Expiration Affects Respiratory Sinus Arrhythmia. Clin Exp Pharmacol Physiol. 2000;27: 601–606. doi:https://doi.org/10.1046/j.1440-1681.2000.03306.x

10. Agelink MW, Malessa R, Baumann B, Majewski T, Akila F, Zeit T, et al. Standardized tests of heart rate variability: normal ranges obtained from 309 healthy humans, and effects of age, gender, and heart rate. Clin Auton Res. 2001;11: 99–108. doi:10.1007/BF02322053

11. Nayak KS, Lim Y, Campbell-Washburn AE, Steeden J. Real-Time Magnetic Resonance Imaging. J Magn Reson Imaging. 2022;55: 81–99. doi:https://doi.org/10.1002/jmri.27411

12. Voit D, Zhang S, Unterberg-Buchwald C, Sohns JM, Lotz J, Frahm J. Real-time cardiovascular magnetic resonance at 1.5 T using balanced SSFP and 40 ms resolution. J Cardiovasc Magn Reson. 2013;15: 79. doi:10.1186/1532-429X-15-79

13. Haji-Valizadeh H, Rahsepar AA, Collins JD, Bassett E, Isakova T, Block T, et al. Validation of highly accelerated real-time cardiac cine MRI with radial k-space sampling and compressed sensing in patients at 1.5T and 3T. Magn Reson Med. 2018;79: 2745–2751. doi:https://doi.org/10.1002/mrm.26918

14. Hauptmann A, Arridge S, Lucka F, Muthurangu V, Steeden JA. Real-time cardiovascular MR with spatio-temporal artifact suppression using deep learning– proof of concept in congenital heart disease. Magn Reson Med. 2019;81: 1143–1156. doi:https://doi.org/10.1002/mrm.27480

15. Piekarski E, Chitiboi T, Ramb R, Feng L, Axel L. Use of self-gated radial cardiovascular magnetic resonance to detect and classify arrhythmias (atrial fibrillation and premature ventricular contraction). J Cardiovasc Magn Reson. 2016;18: 83. doi:10.1186/s12968-016-0306-6

16. Unterberg-Buchwald C, Fasshauer M, Sohns JM, Staab W, Schuster A, Voit D, et al. Real time cardiac MRI and its clinical usefulness in arrhythmias and wall motion abnormalities. J Cardiovasc Magn Reson. 2014;16: P34. doi:10.1186/1532-429X-16-S1-P34

17. Allen BD, Carr ML, Markl M, Zenge MO, Schmidt M, Nadar MS, et al. Accelerated real-time cardiac MRI using iterative sparse SENSE reconstruction: comparing performance in patients with sinus rhythm and atrial fibrillation. Eur Radiol. 2018;28: 3088–3096. doi:10.1007/s00330-017-5283-0

18. Ahmad R, Hu HH, Krishnamurthy R, Krishnamurthy R. Reducing sedation for pediatric body MRI using accelerated and abbreviated imaging protocols. Pediatr Radiol. 2018;48: 37–49. doi:10.1007/s00247-017-3987-6

19. Steeden JA, Kowalik GT, Tann O, Hughes M, Mortensen KH, Muthurangu V. Real-time assessment of right and left ventricular volumes and function in children using high spatiotemporal resolution spiral bSSFP with compressed sensing. J Cardiovasc Magn Reson. 2018;20: 79. doi:10.1186/s12968-018-0500-9

20. Lurz P, Muthurangu V, Schuler PK, Giardini A, Schievano S, Nordmeyer J, et al. Impact of reduction in right ventricular pressure and/or volume overload by percutaneous pulmonary valve implantation on biventricular response to exercise: an exercise stress real-time CMR study. Eur Heart J. 2012;33: 2434–2441. doi:10.1093/eurheartj/ehs200

21. Jakob PM, Griswold MA, Edelman RR, Manning WJ, Sodickson DK. Accelerated Cardiac Imaging Using the SMASH Technique. J Cardiovasc Magn Reson. 1999;1: 153–157. doi:10.3109/10976649909080844

22. Kühl HP, Spuentrup E, Wall A, Franke A, Schröder J, Heussen N, et al. Assessment of Myocardial Function with Interactive Non–Breath-hold Real-time MR Imaging: Comparison with Echocardiography and Breath-hold Cine MR Imaging. Radiology. 2004;231: 198–207. doi:10.1148/radiol.2311021237

23. Bustin A, Fuin N, Botnar RM, Prieto C. From Compressed-Sensing to Artificial Intelligence-Based Cardiac MRI Reconstruction. Front Cardiovasc Med. 2020;7. doi:10.3389/fcvm.2020.00017

24. Baron CA, Dwork N, Pauly JM, Nishimura DG. Rapid compressed sensing reconstruction of 3D non-Cartesian MRI. Magn Reson Med. 2018;79: 2685–2692. doi:https://doi.org/10.1002/mrm.26928

25. Bilal M, Shah JA, Qureshi IM, Kadir K. Respiratory Motion Correction for Compressively Sampled Free Breathing Cardiac MRI Using Smooth l(1)-Norm Approximation. Int J Biomed Imaging. 2018;2018: 7803067. doi:10.1155/2018/7803067

26. Feng L, Grimm R, Block KT, Chandarana H, Kim S, Xu J, et al. Golden-angle radial sparse parallel MRI: combination of compressed sensing, parallel imaging, and golden-angle radial sampling for fast and flexible dynamic volumetric MRI. Magn Reson Med. 2014;72: 707–717. doi:10.1002/mrm.24980

27. Lustig M, Donoho D, Pauly JM. Sparse MRI: The application of compressed sensing for rapid MR imaging. Magn Reson Med. 2007;58: 1182–1195. doi:10.1002/mrm.21391

28. Zhou R, Yang Y, Mathew RC, Mugler JP, Weller DS, Kramer CM, et al. Free-breathing cine imaging with motion-corrected reconstruction at 3T using SPiral Acquisition with Respiratory correction and Cardiac Self-gating (SPARCS). Magn Reson Med. 2019;82: 706–720. doi:10.1002/mrm.27763

29. Wang J, Zhou R, Wang X, Awad M, Salerno M. Free-breathing High-resolution Spiral Real-time Cardiac Cine Imaging at 1.5 T with DEep learning-based Spiral Image REconstruction (DESIRE). ISMRM 30th Annual Scientific Sessions. London, UK: ISMRM; 2022.

30. Ahlander B-M, Maret E, Brudin L, Starck S-A, Engvall J. An echo-planar imaging sequence is superior to a steady-state free precession sequence for visual as well as quantitative assessment of cardiac magnetic resonance stress perfusion. Clin Physiol Funct Imaging. 2017;37: 52–61. doi:https://doi.org/10.1111/cpf.12267

31. Bieri O, Scheffler K. Fundamentals of balanced steady state free precession MRI. J Magn Reson Imaging. 2013;38: 2–11. doi:10.1002/jmri.24163

32. Schär M, Kozerke S, Fischer SE, Boesiger P. Cardiac SSFP imaging at 3 Tesla. Magn Reson Med. 2004;51: 799–806. doi:10.1002/mrm.20024

33. Kellman P, Chefd’hotel C, Lorenz CH, Mancini C, Arai AE, McVeigh ER. Fully automatic, retrospective enhancement of real-time acquired cardiac cine MR images using image-based navigators and respiratory motion-corrected averaging. Magn Reson Med. 2008/02/28. 2008;59: 771–778. doi:10.1002/mrm.21509

34. Larson AC, White RD, Laub G, McVeigh ER, Li D, Simonetti OP. Self-gated cardiac cine MRI. Magn Reson Med. 2004;51: 93–102. doi:https://doi.org/10.1002/mrm.10664

35. Xu J, Van Doren SR. Tracking Equilibrium and Nonequilibrium Shifts in Data with TREND. Biophys J. 2017;112. doi:10.1016/j.bpj.2016.12.018

36. Novillo F, Van Eyndhoven S, Moeyersons J, Bogaert J, Claessen G, La Gerche A, et al. Unsupervised respiratory signal extraction from ungated cardiac magnetic resonance imaging at rest and during exercise. Phys Med Biol. 2019;64: 065001. doi:10.1088/1361-6560/ab02cd

37. Nath MK, Sahambi JS. Independent component analysis of functional MRI data. TENCON 2008 - 2008 IEEE Region 10 Conference. 2008. pp. 1–6. doi:10.1109/TENCON.2008.4766666

38. Jolliffe IT, Cadima J. Principal component analysis: a review and recent developments. Philos Trans R Soc A Math Phys Eng Sci. 2016;374. doi:10.1098/rsta.2015.0202

39. McManus IC, Stöver K, Kim D. Arnheim’s Gestalt Theory of Visual Balance: Examining the Compositional Structure of Art Photographs and Abstract Images. Iperception. 2011;2: 615–647. doi:10.1068/i0445aap

40. Jolliffe IT. Principal Component Analysis. 2nd ed. Springer Series in Statistics. New York: Springer-Verlag; 2002.

41. Calhoun VD, Adali T. Multisubject Independent Component Analysis of fMRI: A Decade of Intrinsic Networks, Default Mode, and Neurodiagnostic Discovery. IEEE Rev Biomed Eng. 2012;5: 60–73. doi:10.1109/RBME.2012.2211076

42. Xu J, Van Doren SR. Tracking Equilibrium and Nonequilibrium Shifts in Data with TREND. Biophys J. 2017;112: 224–233. doi:10.1016/J.BPJ.2016.12.018

43. Du W, Ma S, Fu G-S, Calhoun VD, Adali T. A novel approach for assessing reliability of ICA for FMRI analysis. 2014 IEEE International Conference on Acoustics, Speech and Signal Processing (ICASSP). 2014. pp. 2084–2088. doi:10.1109/ICASSP.2014.6853966

