## Supplementary figures and images for "Simplification of free-running cardiac magnetic resonance by respiratory phase using principal component analysis"

### S1 Fig

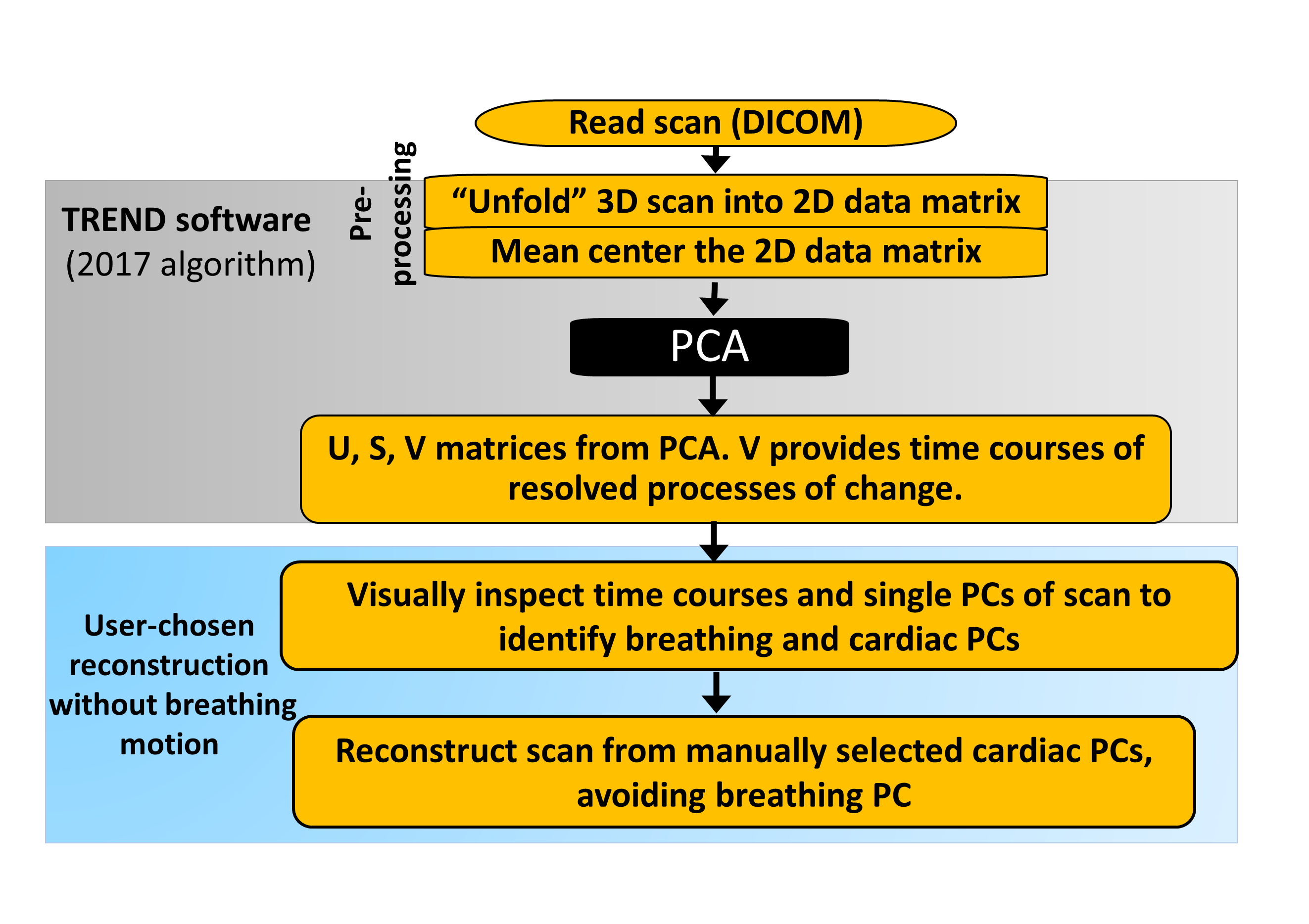

### S2 Fig

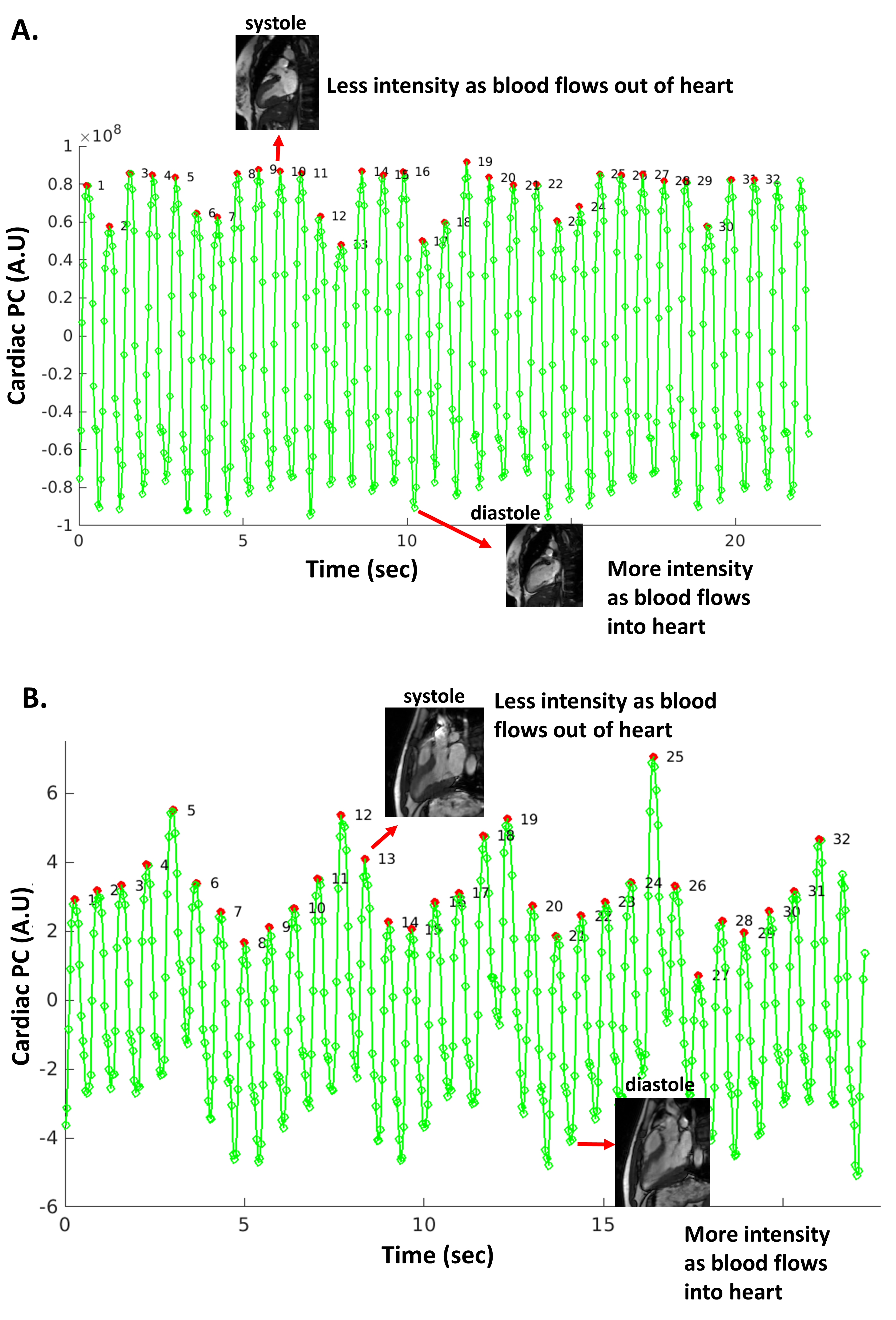

### S3 Fig

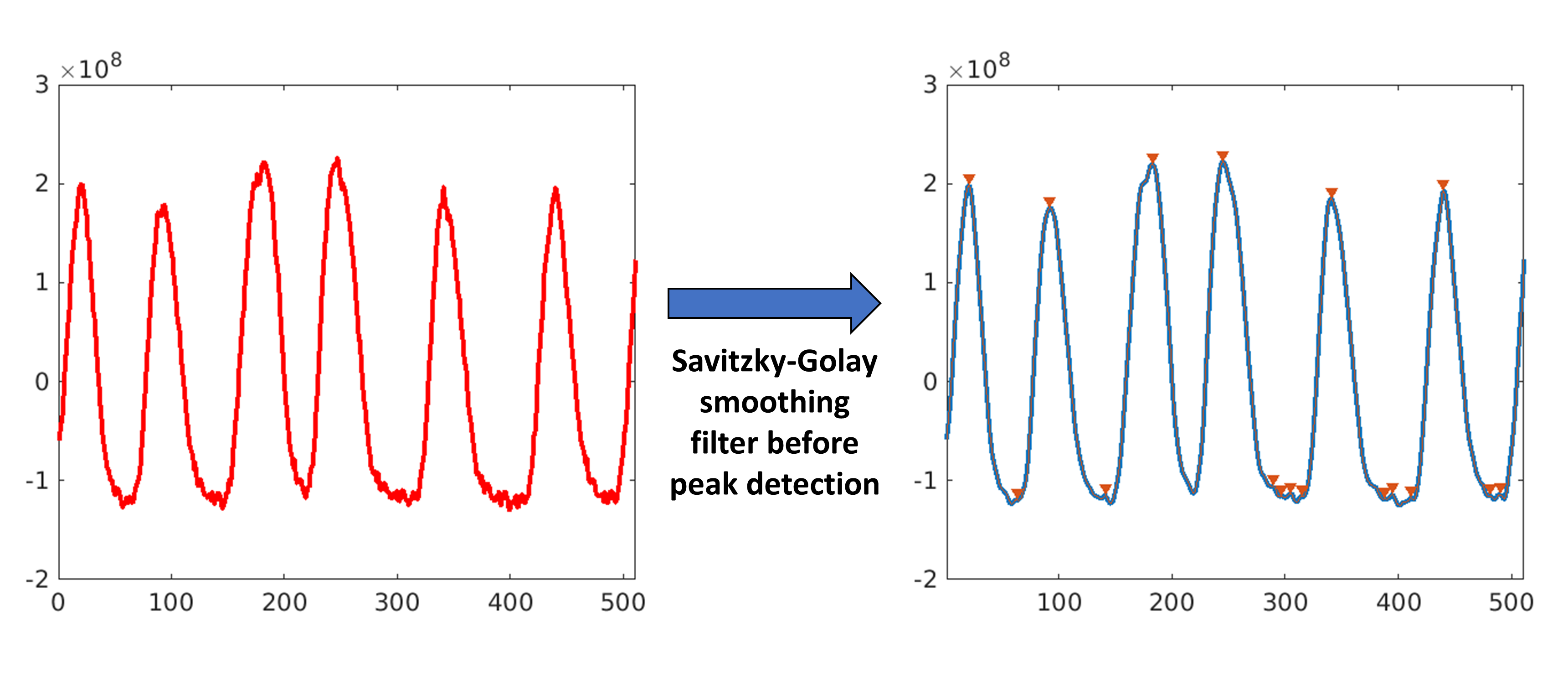

### S4 Fig

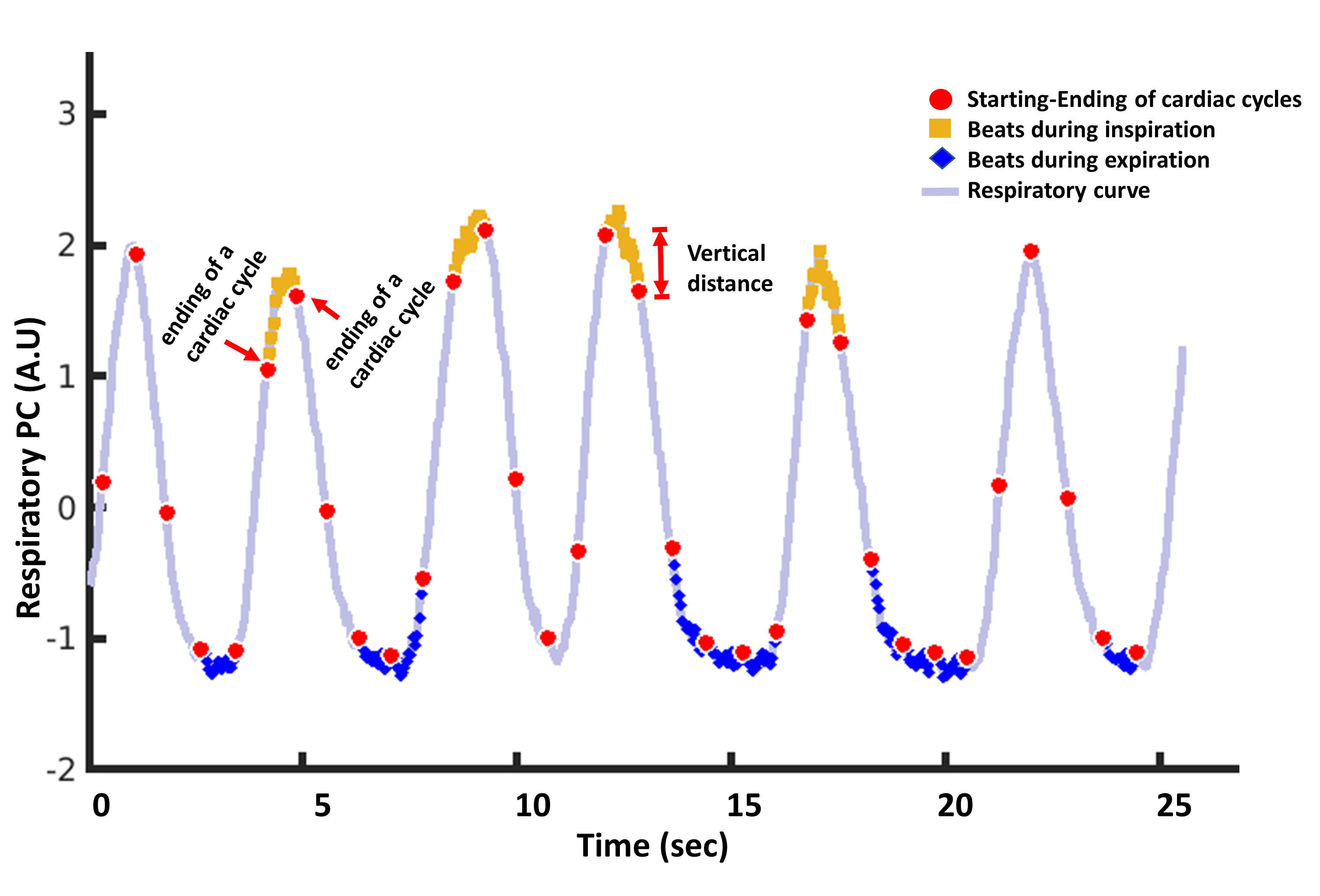

### S5 Fig

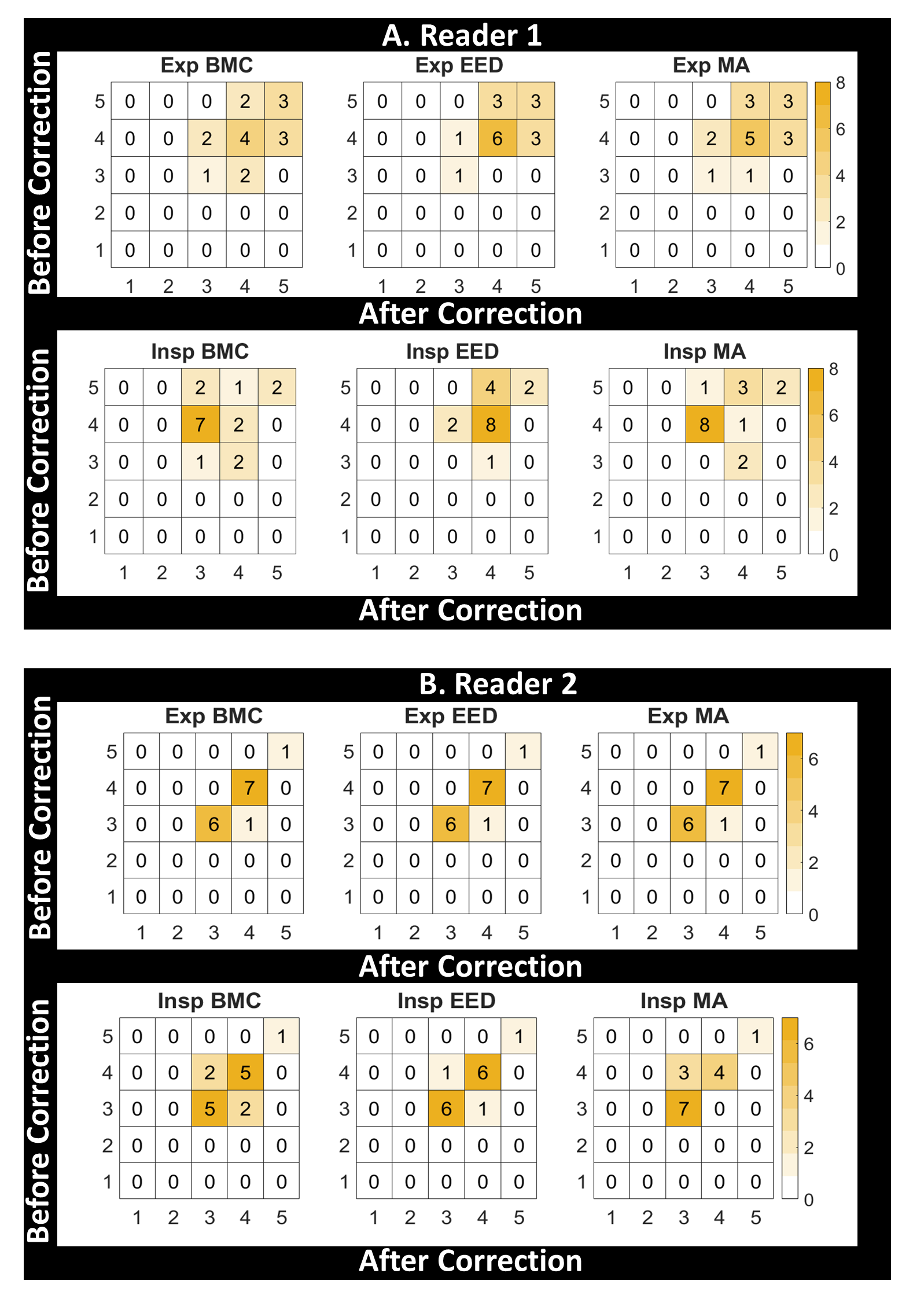
